# Safety and acceptability of clozapine and risperidone in progressive multiple sclerosis: a phase 1, randomized, blinded, placebo-controlled trial

**DOI:** 10.1101/2020.03.12.20034983

**Authors:** Anne Camille La Flamme, David Abernethy, Dalice Sim, Liz Goode, Michelle Lockhart, David Bourke, Imogen Milner, Toni-Marie Garrill, Purwa Joshi, Eloise Watson, Duncan Smyth, Sean Lance, Bronwen Connor

## Abstract

**Objective:** Because clozapine and risperidone have been shown to reduce neuroinflammation in humans and mice, the CRISP trial was conducted to determine whether clozapine and risperidone are suitable for progressive multiple sclerosis (pMS).

**Methods:** The CRISP trial (ACTRN12616000178448) was a blinded, randomized, placebo-controlled trial with three parallel arms (n=12/arm). Participants with pMS were randomized to clozapine (100 to 150 mg/day), risperidone (2 to 3.5 mg/day), or placebo for six months. The primary outcome measures were safety (adverse events/serious adverse events) and acceptability (TSQM-9).

**Results:** An interim analysis (n=9) revealed significant differences in the time-on-trial between treatment groups and placebo (p=0.030 and 0.025, clozapine and risperidone, respectively) with all participants receiving clozapine being withdrawn during the titration period (mean dose=35±15 mg/day). Participants receiving clozapine or risperidone reported a significantly higher rate of adverse events than placebo (p=0.00001) but not serious adverse events. Specifically, low doses of clozapine appeared to cause an acute and dose-related intoxicant effect in patients with pMS, who had fairly severe chronic spastic ataxic gait and worsening over all mobility, which resolved on drug cessation.

**Interpretation:** The CRISP trial results suggest that pMS patients may experience increased sensitivity to clozapine and risperidone and indicate that the dose and/or titration schedule developed for schizophrenia may not be suitable for pMS. While these findings do not negate the potential of these drugs to reduce MS-associated neuroinflammation, they highlight the need for further research to understand the pharmacodynamic profile and effect of clozapine and risperidone in pMS patients.

## INTRODUCTION

Clozapine and risperidone are atypical anti-psychotic agents originally developed in 1959 and 1992, respectively, and are used to treat several neurological disorders including schizophrenia.[1] Both are potent antagonists of a wide range of neuroreceptors including dopamine and serotonin receptors,[1, 2] and recent studies indicate that these agents have anti-inflammatory actions on microglia.[3, 4] Given the known role for neuroinflammation in schizophrenia,[5-7] it is now believed that this class of agents may provide superior benefits during schizophrenia due to their immune-modulatory activity in the CNS.[5, 6, 8-10]

Previous work evaluating the effect of clozapine and risperidone treatment in two mouse models of multiple sclerosis (MS) has demonstrated a dose-dependent reduction in the severity of disease and improvement in disease resolution.[11-13] This protection correlates with reduced CNS inflammation, enhanced functional recovery, and can be achieved at doses that do not promote weight gain, a dose-dependent side effect of both clozapine and risperidone.[12, 13] Together this preclinical work suggests that clozapine and risperidone, which readily pass through an intact blood brain barrier and reduce neuroinflammation in mice and humans,[1] may be useful to treat the chronic neuroinflammation associated with progressive MS (pMS).[14-16]

To this end, the <u>C</u>lozapine and <u>RIS</u>peridone in <u>P</u>rogressive MS (CRISP) trial was designed to investigate to suitability of clozapine and risperidone treatment during pMS. Although there have been several case reports describing the successful use of risperidone or clozapine in MS patients to treat psychosis,[17, 18] this was the first trial of clozapine or risperidone in humans to treat MS itself. The primary aim of this study was to determine if doses of 100-150 mg of clozapine and 2-3.5 mg of risperidone are safe and well-tolerated in patients with pMS.

## RESULTS

Between 8 April 2016 and 8 September 2017, fifty-five people were identified through visits to the Neurology Clinics at Wellington Hospital, hospital databases, regional MS societies, and volunteer recruitment adverts. Of these, twelve were screened at Wellington Hospital for eligibility, and nine enrolled into the CRISP trial and randomly allocated to a treatment arm (Figure 1). There were no significant differences in the baseline clinical parameters between the treatment groups (Table 1) although a significant elevation in WBC and neutrophils (clozapine; p<0.001) and WBC (risperidone; p<0.01) compared to placebo was found (Supplementary Table S1).

**Table 1:** Baseline clinical characteristics.

|  | **Placebo**<br>(n=3) | **Clozapine**<br>(n=3) | **Risperidone**<br>(n=3) |
| --- | --- | --- | --- |
| % Female (n) | 67 (2) | 67 (2) | 100 (3) |
| Age (years) | 58 (12) | 57 (6) | 57 (9) |
| Bodyweight (kg) | 75.8 (21.6) | 89.4 (10.9) | 86.3 (26.6) |
| BMI (kg/m<sup>2</sup>) | 28.4 (3.7) | 30.8 (2.4) | 33.6 (7.9) |
| EDSS (+/- SD) | 6.3 (0.3) | 6.2 (0.6) | 6.2 (0.6) |
| FSS (+/- SD) | 6.4 (0.4) | 4.2 (1.5) | 6.3 (0.8) |
| MSFC (+/- SD) | -1.41 (1.2) | -0.91 (0.2) | -0.76 (1.1) |
| hsCRP (+/- SD) | 3 (0) | 4 (1) | 5.7 (2.3) |
| TropT (+/- SD) | 11.3 (4.7) | 7.3 (4.0) | 5.3 (0.6) |
| Prolactin (+/- SD) | 194 (78) | 203 (87) | 200 (88) |

**Figure 1:**
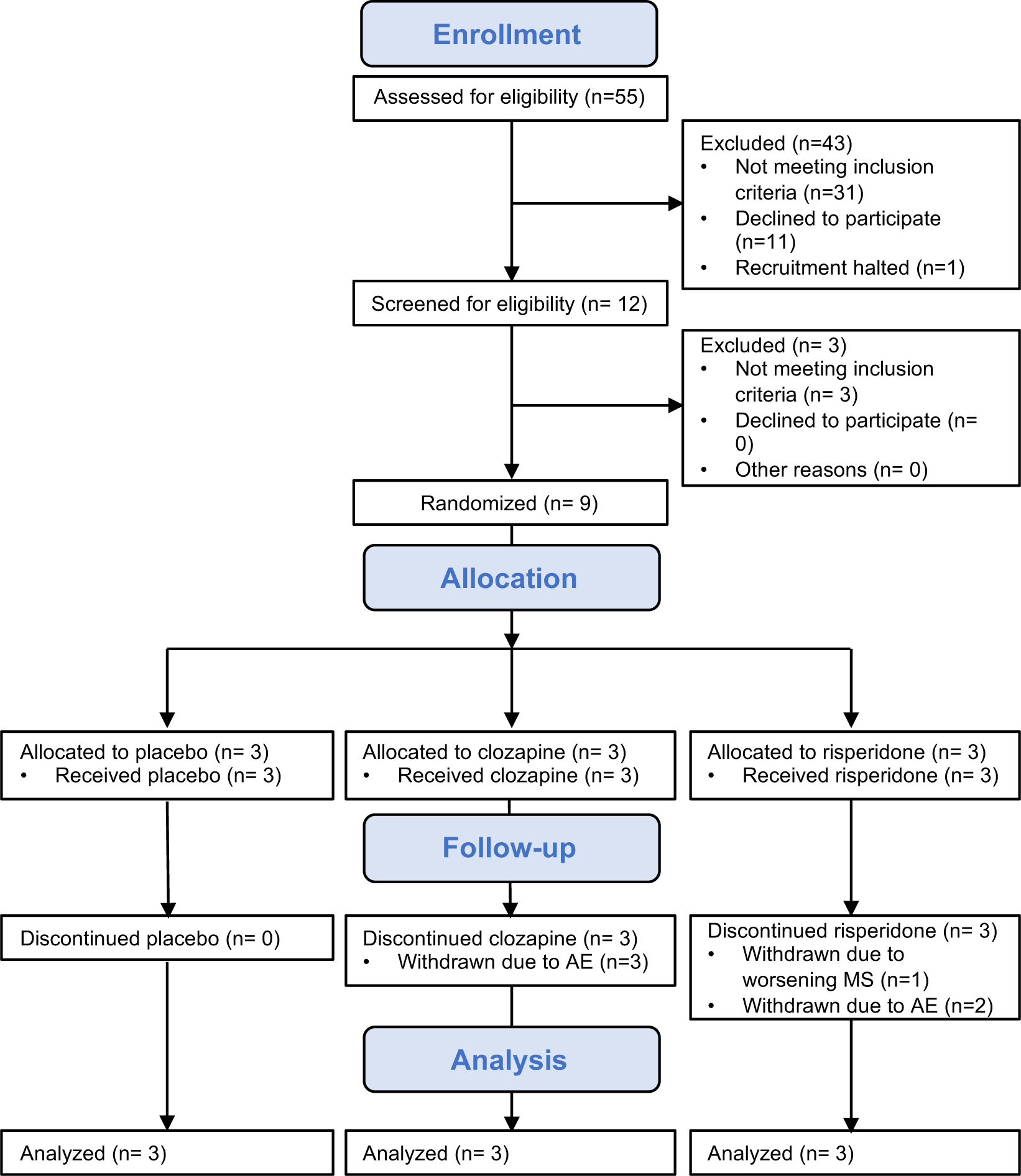
CONSORT diagram for CRISP trial.

Three participants (33%) completed the trial as per protocol while six participants (67%) were withdrawn from the trial; four of whom were withdrawn within the first two weeks. Due to the high number of participants withdrawn from the trial, after completion of enrolment of the first block of nine participants, recruitment was halted for an interim analysis to assess safety (number and rate of AE/SAE) and acceptability (TSQM-9). We used number of the days on trial to compare the total time in the study between each treatment and placebo. The participants who completed the trial (i.e. not withdrawn) were entered as censored observations in the Kaplan-Meier analysis (Figure 2A). We found a significant difference in the time on trial between the groups (p=0.03, P vs C; and p=0.025, P vs R) with a mean time of 8±1 days for participants receiving clozapine and 94±41 days for risperidone compared to 178±3 days for placebo (Figure 2B). Because the doses of the study medications were titrated during the first two weeks when four participants were withdrawn, we compared the percent of the final study medication dose (Figure 2C), which indicated that participants in the clozapine group received 23±10% of the final dose or 35±15 mg/day while those in the risperidone group received 81±33% or 2.8±1.2 mg/day. A comparison of selected baseline parameters between those withdrawn (n=6) and those who completed the trial (n=3), revealed a higher WBC count in those withdrawn (p=0.048 by Wilcoxon test) but no other significant differences (Supplementary Table S2).

**Figure 2:**
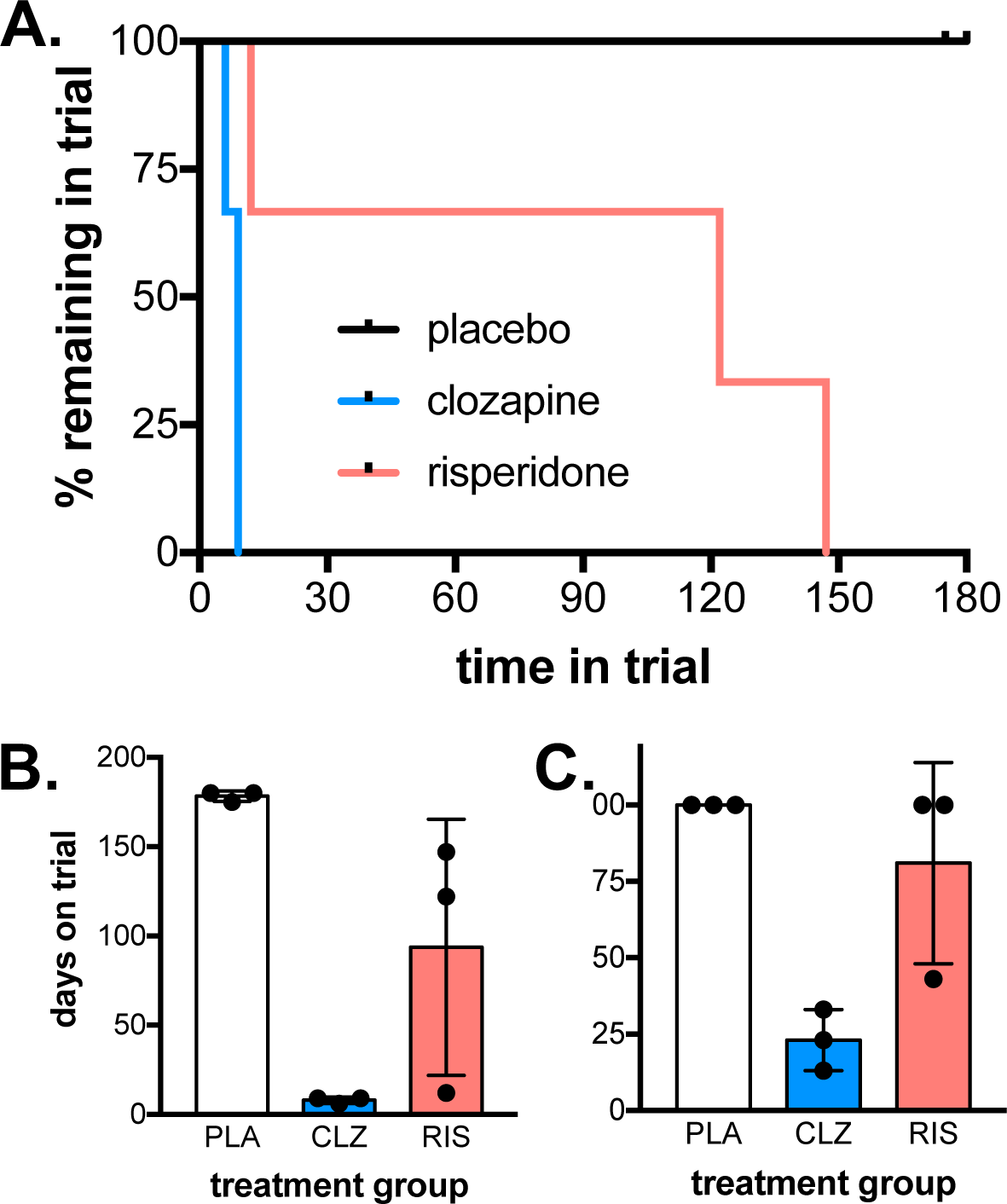
Time on trial was significantly reduced in active treatment arms. (**A**) A Kaplan-Meier plot for the time to withdrawal for each treatment group. CLZ vs PLA (logrank, p = 0.030). RIS vs PLA (p = 0.025). (**B**) Time in days on trial. (**C**) Percentage (%) of final dose at end of study (PLA) or on day of withdrawal (mean CLZ = 35 mg/day, mean RIS = 2.8 mg/day).

The rate of all adverse events was significantly higher (p=0.00001 by Fisher’s exact test) in both clozapine and risperidone groups (37.5 and 11/100 person-days, respectively) compared to placebo (2.43/100 person-days; Table 2). The most common adverse events (≥3 occurrences) assigned a possible, probable, or likely treatment association were drowsiness, dry mouth, dry eyes, muscle weakness, falls, and increased prolactin (Supplementary Table S3). Only three SAE occurred over the whole course of the trial, and no significant differences were found in the rate between treatment groups. One participant receiving risperidone was hospitalized due to rapidly progressing MS. The other two SAE resulted from a participant in the placebo group who broke her right wrist and left clavicle in a motor vehicle accident (Table 2).

**Table 2:** All adverse and serious adverse events.

|  | **Placebo**<br>**(n=3)** | **Clozapine**<br>**(n=3)** | **Risperidone**<br>**(n=3)** |
| --- | --- | --- | --- |
| **CNS** |  |  |  |
| sedation/drowsiness | 0 | 3 | 0 |
| headache | 0 | 0 | 1 |
| parkinsonism | 0 | 0 | 1 |
| balance problems | 1 | 0 | 0 |
| vertigo/dizziness | 1 | 0 | 0 |
| **Gastrointestinal** |  |  |  |
| dry mouth | 0 | 2 | 3 |
| hypersalivation | 0 | 0 | 1 |
| constipation | 1 | 0 | 0 |
| nausea | 1 | 0 | 0 |
| **Neuromuscular** |  |  |  |
| rapid progression of weakness | 0 | 0 | 1* |
| muscle weakness | 0 | 3 | 2 |
| leg dragging | 1 | 0 | 0 |
| **Other** |  |  |  |
| dry eyes | 0 | 0 | 3 |
| rash | 0 | 0 | 1 |
| fall | 0 | 1 | 13 |
| increased prolactin | 0 | 0 | 2 |
| pain or aches | 2 | 0 | 1 |
| itchiness | 1 | 0 | 0 |
| can't stand smell of meat | 1 | 0 | 0 |
| sore throat | 1 | 0 | 0 |
| fatigue | 0 | 0 | 1 |
| vivid dreams | 0 | 0 | 1 |
| urinary tract infection | 1 | 0 | 0 |
| broken wrist, broken clavicle<br>(motor vehicle accident) | 2* | 0 | 0 |
| **TOTAL** | 13 | 9 | 31 |
| # of person-days | 535 | 24 | 281 |
| Normalized (per 100 person-days) | 2.43 | 37.5 | 11 |
| P value** |  | 0.00001 | 0.00001 |

\* Serious adverse event
\*\* Calculated by Fisher's exact test compared to placebo.

For those participants who completed at least three months of treatment (n=3, placebo and n= 2, risperidone), no difference in treatment acceptability was found in any of the three domains (efficacy, convenience, and global satisfaction) assessed by the TSQM-9 (Table 3). Additionally, after three months of treatment, there were no significant changes from baseline parameters including bodyweight, BMI, EDSS, FSS, MSFC, WBC, neutrophils, TropT, and CRP except for an elevation in serum prolactin in the risperidone group as expected (Table 4; Supplementary Figure S1).[19]

**Table 3:** Treatment satisfaction questionnaire for medicine – 9 (TSQM-9) at 3 months.

|  | **Efficacy** | **Convenience** | **Global satisfaction** |
| --- | --- | --- | --- |
| **Placebo (n=3)** |  |  |  |
| Mean (SD) | 66.7 (17) | 77.8 (25) | 71.4 (25) |
| Median (min, max) | 66.7 (50, 83.3) | 83.3 (50, 100) | 57.1 (57.1, 100) |
| **Risperidone (n=2)** |  |  |  |
| Mean (SD) | 58.4 (4) | 94.5 (8) | 42.9 (20) |
| Median (min, max) | 58.4 (55.6, 61.1) | 94.5 (88.9, 100) | 42.9 (28.6, 57.1) |
| P value* | 0.564 | 0.374 | 0.197 |

\* Calculated by Kruskal-Wallis test to compare means between groups.

**Table 4:** Changes to clinical parameters after risperidone treatment – 3 months.

| **Parameter:**<br>**mean (SD)** | **Time**<br>**(months)** | **Placebo**<br>**(n=3)** | **Risperidone**<br>**(n=2)** |
| --- | --- | --- | --- |
| Bodyweight (kg) | 0 | 75.9 (21) | 71.2 (6) |
|  | 3 | 75.8 (21) | 69.5 (3) |
| BMI | 0 | 28.4 (4) | 29.3 (4) |
|  | 3 | 28.4 (4) | 28.6 (3) |
| EDSS | 0 | 6.3 (0.3) | 6.0 (0.7) |
|  | 3 | 6.2 (0.3) | 6.3 (1.0) |
| FSS | 0 | 6.4 (0.4) | 6.1 (1.0) |
|  | 3 | 6.1 (0.2) | 5.6 (1.0) |
| MSFC | 0 | -1.4 (1.2) | -1.1 (1.2) |
|  | 3 | -1.1 (0.4) | -1.3 (1.1) |
| WBC | 0 | 5.9 (1.1) | 7.5 (1.3) |
|  | 3 | 5.7 (0.9) | 6.9 (0.1) |
| Neutrophils | 0 | 3.7 (0.3) | 4.7 (1.4) |
|  | 3 | 3.6 (0.2) | 4.5 (1.1) |
| TropT | 0 | 11.3 (4.7) | 5.5 (0.7) |
|  | 3 | 13.0 (5.3) | 5.0 (0.0) |
| CRP | 0 | 3.0 (0.1) | 5.0 (2.8) |
|  | 3 | 3.0 (0.1) | 4.5 (2.1) |
| Prolactin – ng/mL | 0 | 194 (78) | 216 (117) |
|  | 3 | 271 (154) | 4207 (1854) |

## DISCUSSION

This study is the first report of the use of clozapine and risperidone to treat pMS and revealed an unexpected sensitivity in this population to clozapine and, to a lesser extent, risperidone. Specifically, all participants receiving active treatment were withdrawn from the study with the clozapine-treated group averaging only eight days while the risperidone-treated participants remained in the trial for an average of ninety-four days. In comparison, all participants in the placebo group completed the trial as per protocol. The decreased time in trial was associated with an increased rate of AE but not SAE with the key AE being dry mouth, drowsiness, dry eyes, muscle weakness, and falls. This increased sensitivity to clozapine as well as risperidone in people with pMS is particularly unexpected given that the doses administered were exceptionally low for clozapine (35 mg/day at study termination) and moderate to low for risperidone (2.8 mg/day).[20-23]

When initiating atypical anti-psychotic treatment, a slow titration is recommended over a two-week period until the therapeutic dose is reached.[24, 25] For schizophrenia, this dose is 350-400 mg/day for clozapine and 2-6 mg/day for risperidone.[21, 22] The titration allows a slow tolerization to the drug and reduces side effects.[1] For clozapine, a dose of 12.5 mg/day is recommended for the first dose when used for schizophrenia or Parkinson’s disease.[24] In contrast, the starting clozapine dose in the CRISP trial was 5 mg/day allowing a very slow and conservative titration up to the target dose (100 mg/day) for the first three months. Despite the low dose, adverse events occurred by day three, equating to 15 mg/day, and participants were withdrawn at a mean dose of 35 mg/day. While 7.1 to 15.6% of patients may be withdrawn from clozapine due to adverse events,[20] this is the first report of a categorical sensitivity to clozapine at extremely low doses by a cohort of individuals.

During clozapine titration, a common adverse event is orthostatic hypotension, and this hypotension occurs even at a dose of 12.5 mg/day.[26] Although we did not test specifically for orthostatic hypotension due to the progressive MS disability, no effect on blood pressure was detected at any time during the four-hour monitoring period post-doses one and two of clozapine (Supplementary Figure S2). In addition to the initial orthostatic hypotension, the most common AE occurring in more than one in ten patients administered clozapine are weight gain, drowsiness, sedation, dizziness, tachycardia, constipation, and hypersalivation.[21, 26] For the CRISP participants, all three patients allocated to clozapine developed a similar syndrome with lower limb weakness alone in one, lower limb weakness and poor balance in the second, and drowsiness followed by poor balance in the third. In the first two, it began at day five (35 mg) and worsened over four days (35-50 mg) until they could no longer walk. The third noted drowsiness after the third dose (15 mg) and unsteadiness the following day after 20 mg. Medication was stopped. She was worse the following day with a fizzy feeling in her fingers, an exacerbation of a pre-existing symptom, but recovered a day later. Rechallenge with a lower dose produced identical symptoms. In all three cases, the symptoms wore off within forty-eight hours and resembled an intoxication. Their EDSS scores at study entry were 6.5, 6.5, and 5.5, respectively. These observations suggest that patients with advanced MS with cerebellar and pyramidal tract dysfunction may be unusually susceptible to intoxication by clozapine.

Although two participants experienced few adverse events during the risperidone titration period, one participant experienced rashes after the first dose, treatment was halted, and then rechallenged one week later. However, the participant was unable to tolerate risperidone and was withdrawn. Adverse cutaneous reactions occur in 2-5% of individuals treated with psychiatric medications including atypical antipsychotics and predominantly occur within the first two weeks.[27, 28] Exanthematous rashes have been associated with risperidone and resolve after discontinuing treatment.[29]

Other AE experienced by the risperidone-treated group included dry mouth, dry eyes, muscle weakness, falls, and increased prolactin. Whereas dry eyes and dry mouth are common (1-10%) adverse events, muscle weakness is far less common (0.1-1%). Risperidone is associated with elevated serum prolactin and by three months, the risperidone group had elevated levels of prolactin as expected.[19] Interestingly, one side effect that was not observed in the risperidone group was weight gain, which has been reported in one study to occur in 58.4% of patients after three months (>7% gain).[22, 30]

In conclusion, we report an increased sensitivity to clozapine and risperidone in a pMS cohort. Furthermore, the sensitivity is striking in that 1) it occurred at very low doses for clozapine and 2) the adverse reaction pattern was different from expected with no occurrence of hypotension or weight gain and increased reports of muscle weakness and falls. However, despite having statistically significant findings, this study has limitations that need to be considered, the most important of which is the very low number of participants in each treatment group. Although possible that our findings are within the expected adverse event pattern, the categorical early withdrawal of *all* participants from the active arms provides strength to our conclusions.[20] An additional limitation is that with the unexpected withdrawal of 44% of the participants within the first two weeks, no samples were collected to confirm the serum levels of the study medication in these subjects. Finally, because this study was not powered to assess efficacy, the potential of these two atypical antipsychotics to treat the neuroinflammation associated with pMS, was not evaluated. However, current studies are underway to assess the safety and tolerability of quetiapine (300 mg daily), a clozapine analogue, as a remyelinating therapy in MS.[31, 32] Given the results of our trial using low doses of clozapine, our findings suggest that it may not be appropriate to administer either clozapine or risperidone in the same manner during pMS as during schizophrenia or the other neurological disorders for which they are indicated.

## METHODS

### Study design and participants

The CRISP study was a blinded, placebo-controlled trial of clozapine and risperidone treatment in pMS patients conducted at Wellington Regional Hospital in Wellington, New Zealand. Initially, the trial planned to recruit thirty-six participants (n = 12/group) to receive clozapine, risperidone, or placebo for six months. An interim analysis was performed after the first block of nine participants, and here we report the results from that interim analysis. Participants were identified and recruited from the neurology ward, neurology outpatient clinics, community MS support group meetings, and after a review of existing databases. We recruited nine participants that fulfilled the inclusion criteria of a diagnosis of pMS with the continuous worsening of neurological impairment over at least six months, aged 18-70 years, and an Expanded Disability Status Scale (EDSS) of 3.5-7.0. Full inclusion criteria can be found in the Supplementary Information.

Participants were excluded if they were or had a current diagnosis of RRMS; pregnant or lactating; unable to undergo regular blood tests or MRI scans; contraindications to clozapine or risperidone; known hypersensitivity to clozapine, risperidone or to any of the excipients thereof; past intolerance to clozapine or risperidone; *c*oncomitant use of medications known to affect clozapine treatment; concomitant disease likely to interfere with the trial medication; serious medical co-morbid illness or any other disease or condition which, in the opinion of the investigator, means that it would not be in the patient’s best interests to participate in the study. The full exclusion criteria can be found in the Supplementary Information.

### Standard protocol approvals, registrations, and patient consents

The CRISP study was approved by the New Zealand Central Health and Disability Ethics Committee (15/CEN/216) and the Standing Committee on Therapeutic Trials (15/SCOTT/177), and the trial was registered (11/02/2016) in the Australian New Zealand Clinical Trials registry (ACTRN12616000178448). All participants provided written informed consent prior to screening. Additionally, all methods were performed in accordance with the relevant guidelines and regulations.

### Randomization and blinding

Permuted-block randomization using a block size of nine with no stratification was generated using the statistical software R to allocate a study medication to each participant study number. The treating neurologist, assessing neurologist, research nurse, and the participants were blinded as to the treatment allocation. For the interim analysis, the data bases were locked after review by the trial site staff and independent trial monitor. The trial statistician then analyzed the primary and secondary outcome measures from the interim data.

### Procedures

Participants provided written consent and then were screened. Baseline measurements collected including EDSS, MSFC, FSS, EEG, and blood tests, and if eligible, participants were enrolled and randomised into the study. Participants received clozapine suspension (50 mg/ml), risperidone tablets (0.5 mg, 2.0 mg, and 3.0 mg tablets) or placebo suspension (matched to clozapine suspension); study medications were provided by Douglas Pharmaceuticals (Auckland, New Zealand). The study medication was titrated slowly over a two-week period starting at 5 mg/day (clozapine) and 0.5 mg/day risperidone until participants reached 100 mg/day clozapine and 2 mg/day risperidone (Supplementary Table S4). After 2.5 months, the dose of study medication was increased over a two-week period up to 150 mg/day and 3.5 mg/day of clozapine and risperidone, respectively (Supplementary Table S4). Participants were monitored in Wellington Hospital for four hours after each of the first five doses to assess body temperature, blood pressure lying, heart rate, and adverse events as per recommended guidelines for clozapine treatment.[24, 33] After six months, participants were titrated off the study medication over a two week period and four weeks later, had their final end of study visit.

### Outcomes

Participants visited Wellington Hospital or were contacted by the research nurse by phone on alternate weeks to assess adverse events and monitor compliance. Additionally, the treating neurologists assessed adverse and serious adverse events during the baseline, three month, and six month visits and throughout the trial as required.[20, 34] Full blood counts (FBC) were assessed at baseline, weekly for the first eighteen weeks and every four weeks thereafter until week 26 (six-month visit). A final FBC was taken at week 32 (end of study). Alanine aminotransferase, alkaline phosphatase, gamma glutamyltransferase, total bilirubin, serum albumin, total serum protein, serum creatinine, creatine phosphokinase, glycosylated haemoglobin, and prolactin were assessed at baseline, three-months, and six-months. C reactive protein and troponin T were assessed at baseline, weekly for the first four weeks, and at three- and six-months. The full schedule of haematological parameters can be found in Supplementary Table S5. MRI was assessed at baseline and six months by Pacific Radiology (Wellington, New Zealand) and an echocardiogram was performed at baseline by Wakefield Cardiology (Wellington, New Zealand). At baseline, three months, and six months, the EDSS, MSFC, FSS, and Treatment Satisfaction for Medicines Questionnaire-9[35] (TSQM-9; three- and six-months only) were administered by separate, blinded assessors.

### Statistical analyses

The statistical analyses were designed and conducted by DS. For the primary outcome measures, the rates of AE/SAEs were compared between groups using the number of events per patient-week using OpenEpi software and Fisher’s exact tests. The total time in the study between the three groups was compared using Kaplan-Meier analysis (Survival analysis) and the logrank test. Finally, the TSQM-9 at 3-months was compared between groups using a Kruskal-Wallis test. For secondary analyses, to determine if there was any difference between the participants who withdrew or were withdrawn from the study and those who did not withdraw/were not withdrawn, baseline data on all participants including clinical parameters (EDSS, age, and hematological parameters) were compared by Wilcoxon tests.

Post-hoc analyses to assess differences in clinical parameters and leukocyte populations between treatment groups at baseline were also analyzed using Kruskal-Wallis (for three groups) or Wilcoxon (for two groups) so that we did not have to assume the parameters were normally distributed. Post-hoc analyses are indicated in the table legends and were performed using GraphPad Prism (La Jolla, CA, USA) software version 7.

## Data Availability

Deidentified data collected during this study and presented in this manuscript are available from the corresponding author on request from individuals affiliated with research or health care institutions.

## ACKNOWLEDGEMENTS

The authors would like to thank the staff at the Neurology Department and the Clinical Trials Unit at Wellington Regional Hospital for their support throughout the CRISP trial, and in particular, Marina Dzhelali, Jonathan Barrett, and Angela McDonnell. Finally, the authors appreciate the advice and help provided by Dr Lisa Woods (Victoria University of Wellington) in generating the randomization sequence.

## AUTHOR CONTRIBUTIONS

The authors contributed to the study as follows: study design (ACL, DA, DS, BC), data collection (DA, LG, DB, IM, TG, PJ, EW, DS, SL), data analysis and interpretation (ACL, DA), statistical analysis (DS), independent trial monitoring (ML), writing (ACL), and editing (DA, DS, BC). ACL, DA, LG, ML, and DB had full access to the data, reviewed manuscript drafts, and approved the final manuscript. ACL had final responsibility for the decision to submit for publication.

